# VMAP: Vaginal Microbiome Atlas During Pregnancy

**DOI:** 10.1101/2023.03.21.23286947

**Authors:** Antonio Parraga-Leo, Tomiko T. Oskotsky, Boris Oskotsky, Camilla Wibrand, Alennie Roldan, Alice Tang, Connie W.Y. Ha, Ronald J. Wong, Samuel S. Minot, Gaia Andreoletti, Idit Kosti, Kevin R. Theis, Sherrianne Ng, Yun S. Lee, Patricia Diaz-Gimeno, Phillip R. Bennett, David A. MacIntyre, Susan V. Lynch, Roberto Romero, Adi L. Tarca, David K. Stevenson, Nima Aghaeepour, Jonathan Golob, Marina Sirota

## Abstract

The vaginal microbiome has been shown to be associated with pregnancy outcomes including preterm birth (PTB) risk. Here we present VMAP: Vaginal Microbiome Atlas during Pregnancy (http://vmapapp.org), an application to visualize features of 3,909 vaginal microbiome samples of 1,416 pregnant individuals from 11 studies, aggregated from raw public and newly generated sequences via an open-source tool, MaLiAmPi. Our visualization tool (http://vmapapp.org) includes microbial features such as various measures of diversity, VALENCIA community state types (CST), and composition (via phylotypes and taxonomy). This work serves as a resource for the research community to further analyze and visualize vaginal microbiome data in order to better understand both healthy term pregnancies and those associated with adverse outcomes.

## Introduction

The vaginal microbiome plays a significant role in birth outcomes, including spontaneous and preterm labor^1–7^. An increasing number of microbiome studies in the past decade, thanks to the advances in high-throughput sequencing, have permitted greater insights into the role of the microbiome in human health and disease^8^; however, efforts to aggregate microbiome datasets to better understand the microbiome through analysis of large, widely-representative data has been precluded by technical challenges^9^. In this work, we present an atlas of the vaginal microbiota during pregnancy leveraging data across 11 studies harmonized by the open-source tool, MaLiAmPi. This dataset represents one of the largest and most geographically diverse aggregations of the vaginal microbiome in pregnancy to date – 3,909 samples from 1,416 pregnant individuals – harmonized into a set of generalizable features suitable for integration of new microbiota data post-hoc. A rich set of paired metadata is included, including collection week during gestation (by specimen), week of delivery, and maternal NIH racial category. Approximately one-third of the pregnancies in this data set were of women delivering preterm, including just over 1/8th who delivered early preterm. This dataset and features derived from these data can be interactively visualized via our Vaginal Microbiome Atlas During Pregnancy (VMAP) application, a resource for understanding the vaginal microbiome in the context of pregnancy (http://vmapapp.org).

## Methods

The dataset was constructed by aggregating and processing vaginal microbiome data from the public domain^10–14^ in addition to including newly generated data not yet released to the public. The publicly available data were from nine studies, representing 3,578 samples across 1,268 individuals of whom 851 delivered term and 417 preterm (before 37 weeks of gestation) including 170 whose deliveries were early preterm (before 32 weeks of gestation). Two additional unpublished datasets are included. One is from Wayne State University consisting of 159 samples across 60 individuals among whom 40 (66.7%) had term deliveries and 20 (33.3%) had preterm deliveries, including 5 (8.3%) who had early preterm deliveries. A newly generated dataset which comprises 172 vaginal microbiome samples from 88 individuals, up to three samples (one sample per trimester) for each individual, with 48 individuals (54.5%) having term deliveries, and 40 individuals (45.5%) having preterm deliveries including 8 (9.1%) having early preterm deliveries^15^.

To harmonize the raw amplicon sequence reads from such a diverse set of underlying approaches, we developed a novel phylogenetic-based approach implemented in an open source workflow called MaLiAmPi^14^. We further compute the alpha-diversity of communities (diversity measures include Shannon, Inverse Simpson, Balance weighted phylogenetic diversity (bwpd), phylogenetic entropy, quadratic, unrooted phylogenetic diversity, and rooted phylogenetic diversity), weighted phylogenetic (KR) distance between communities, provide taxonomic assignments to each ASV, and cluster ASVs into phylotypes which serve as a taxonomy-independent representative of (phylogenetically) closely related organisms. In addition, VALENCIA^17^ was used to provide the community state type (CST) of each sample. VMAP, an interactive R Shiny application was developed to visualize the data.

## Results

VMAP interactively visualizes vaginal microbiome features during healthy term or adverse-outcome associated pregnancies across 11 studies, representing 3,909 samples from 1,416 individuals, of whom 939 delivered at term and 477 preterm, including 183 whose deliveries were early preterm. The demographics and information about the source studies are presented in Table 1. A screen shot of the visualization tool is presented in Figure 1. This tool allows customizable visualization of the data at several levels including demographics of the cohort at the sample or individual level organized by outcome (term, preterm, early preterm), race and ethnicity distributions, as well as breakdown per project. Demographics are shown as stacked barplots where the relative frequency of each type of category selected can be seen. Alpha-diversity measures are also shown in several ways including overall correlation plots of the individual measures and their distribution as well as box plots across various stratifications. Different diversity measures are customizable allowing users to select their measures of interest. CSTs are visualized longitudinally across trimesters allowing for various stratifications of interest. This kind of plot allows the observation of changes in the composition of CSTs across trimesters according to the feature selected, representing each stream as an individual. Finally, dimensionality reduction plots, namely Uniform Manifold Approximation and Projections (UMAPs), are used to visualize phylotype data per sample or individual comparing among different features. The prevalence and relative abundance of closely related microbes (as represented by phylotypes) or taxa are displayed as heatmaps (for prevalence).

**Table 1:** Summary of studies, individuals, samples, and 16S rRNA gene sequencing information

|  |  |  |
| --- | --- | --- |
| Studies | Accession IDs | SDY465, PRJEB11895, PRJEB21325, PRJEB30642, PRJNA242473, PRJNA294119, PRJNA393472, PRJNA430482, PRJNA504518, PRJEB12577 |
|  | Centers | Imperial College London, Stanford University, University of Maryland, University of Pennsylvania, Virginia Commonwealth, Washington University, Wayne State University |
| Individuals | n | 1416 |
| Age Range, n (%) | Unknown | 691 (48.8) |
|  | Below 18 | 4 (0.3) |
|  | 18 to 28 | 304 (21.5) |
| Race, n (%) | 28 to 38 | 357 (25.2) |
|  | Above 38 | 60 (4.2) |
|  | Race: American Indian or Alaska Native | 9 (0.6) |
|  | Race: Asian | 84 (5.9) |
|  | Race: Black or African American | 827 (58.4) |
|  | Race: Native Hawaiian or Other Pacific Islander | 7 (0.5) |
|  | Race: White | 422 (29.8) |
|  | Race: Unknown | 71 (5.0) |
| Ethnicity, n (%) | Ethnicity: Hispanic or Latino | 50 (3.5) |
|  | Ethnicity: Unknown | 1261 (89.1) |
| Delivery, n (%) | Term | 939 (66.3) |
|  | Preterm | 477 (33.7) |
|  | Early Preterm | 183 (12.9) |
| Samples | n | 3909 |
| Delivery, n (%) | Term | 2786 (71.3) |
|  | Preterm | 1123 (28.7) |
|  | Early Preterm | 363 (9.3) |
| 16S rRNA Sequencing | V Region Sequences | V1 - V2, V1 - V3, V3 - V4, V3 - V5, or V4 |
|  | Instruments | 454 GS FLX Titanium, Illumina HiSeq 2500, Illumina HiSeq 4000, Illumina MiSeq, or Illumina NextSeq 550 |

**Figure 1:**
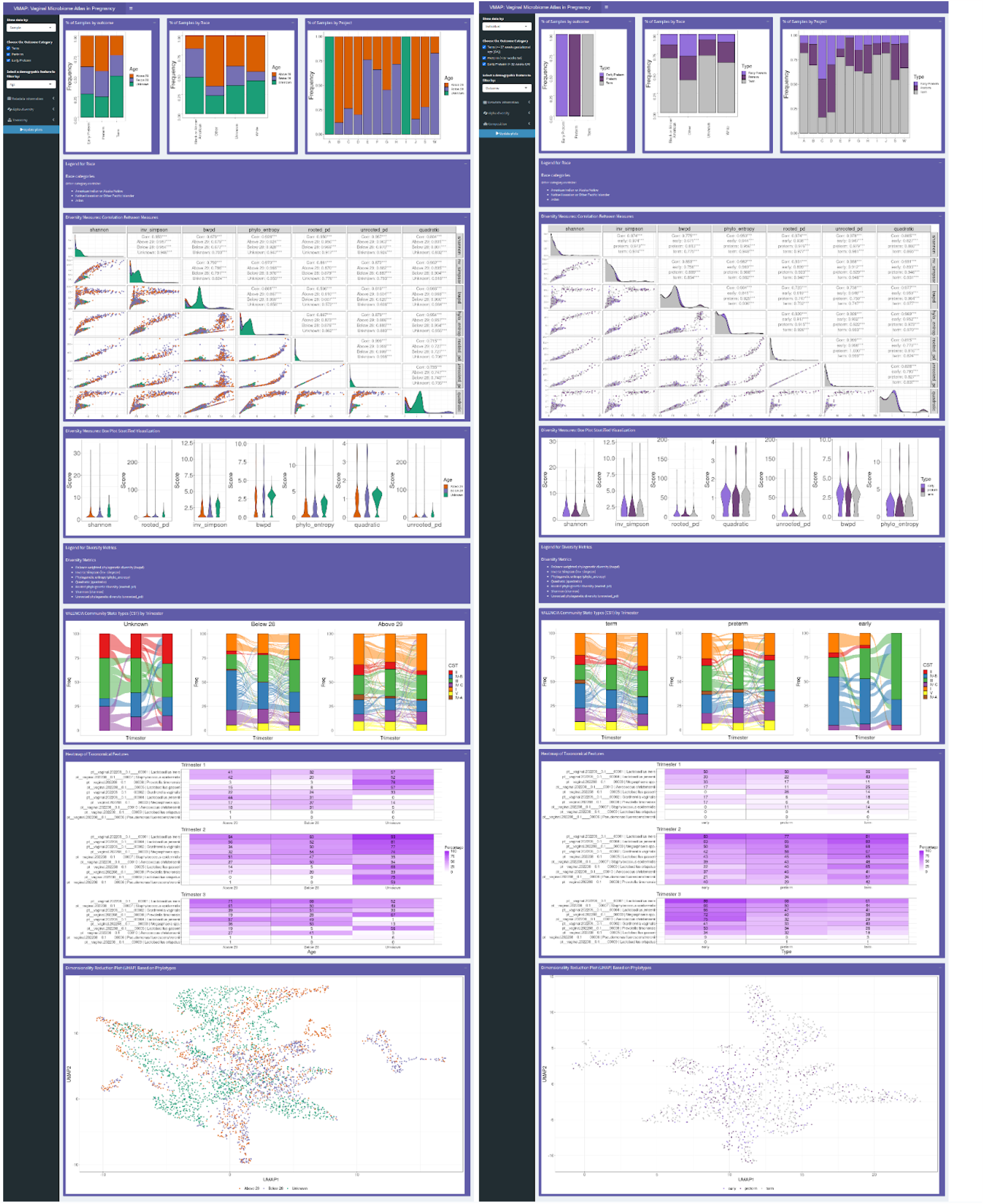
VMAP RShiny App Screenshot. A user can visualize demographic features of the cohort, diversity measures (both correlation between them) and each one by outcome of interest, CSTs as alluvial plots, dimensionality reduction plots of phylogeny and heatmaps of taxonomical features. The visualizations are customizable by the bar on the left allowing one to choose which projects and samples to visualize. A) visualized by age B) visualized by outcome

## Discussion

VMAP provides a means to view a rich set of features derived from vaginal microbiota data (including alpha-diversity, CSTs, and the composition of specific microbes and taxa) associated with core outcomes (delivery week) and metadata (e.g., NIH racial categories, maternal age) that can be sliced and viewed interactively. VMAP can be a resource for those wishing to relate and understand vaginal microbiota data within the context of pregnancy. The generalizability of these features across studies using distinct underlying techniques (e.g., targeting different 16S rRNA gene variable regions or novel sequencing platforms) and the ability to integrate new data into the existing set of features are significant advances for microbiome research. For example, this data set and approach were successfully employed as the basis for a machine learning (“DREAM”) challenge predicting preterm and early-preterm birth where over 300 teams participated with the goal of building machine learning models to predict which women would deliver preterm^15^. This challenge used this novel approach to integrate data *post-hoc* to validate machine learning models against datasets unavailable to participants.

The resource has limitations that should be considered. It is based on publicly available data which might not have full clinical or demographic annotations of the samples in the metadata. While the sample size of the study is considerable, it may not be representative of the entire population of pregnant women from around the world.

This work serves as the basis for several potential follow-up opportunities. Our visualization resource can further be extended to include non-pregnancy datasets as well as microbiome data across body sites. In addition, other adverse pregnancy outcomes such as recurrent pregnancy loss can be investigated and included in the expansion of the resource. Finally, the microbiome data can further be integrated with other omics measures to better understand healthy human pregnancy and those associated with adverse outcomes. VMAP will be valuable for more robust interpretation of novel datasets seeking to relate the vaginal microbiome to pregnancy outcomes – serving as a large and regularized set of data and metadata for comparison.

## Code and Data Availability

The RShiny app is available: http://vmapapp.org

Datasets were downloaded from ImmPort^11^ via the March of Dimes Preterm Birth database^10^ (Study SDY465), from the NCBI Sequence Read Archive^13^ (BioProjects PRJNA242473, PRJNA294119, PRJNA393472, and PRJNA430482), the Sequence Read Archive of the European Nucleotide Archive^14^ (Projects PRJEB11895, PRJEB12577, PRJEB21325, and PRJEB30642), and the database of Genotypes and Phenotypes (dbGaP)^12^ (accession number phs001739.v1.p1).

Additional associated metadata were requested and obtained from the RAMS Registry (https://ramsregistry.vcu.edu) (PRJNA430482) or from the senior author (Projects PRJEB11895, PRJEB12577, PRJEB21325, and PRJEB30642), or downloaded from the publication by the Kindinger et al.(ref) (PRJEB11895 and PRJEB12577).

Data for accession number phs001739.v1.p1 are exclusively available via dbGap after following the application procedures there.

The aggregated vaginal microbiome dataset can be downloaded from the March of Dimes Prematurity Research Database excluding the datasets that we are unable to share due to legal restrictions (**SDY2187**).

The code for VMAP is available on Github: https://github.com/msirota/vmap.git

## Data Availability

The RShiny app is available: http://vmapapp.org
Datasets were downloaded from ImmPort11 via the March of Dimes Preterm Birth database10 (Study SDY465), from the NCBI Sequence Read Archive13 (BioProjects PRJNA242473, PRJNA294119, PRJNA393472, and PRJNA430482), the Sequence Read Archive of the European Nucleotide Archive14 (Projects PRJEB11895, PRJEB12577, PRJEB21325, and PRJEB30642), and the database of Genotypes and Phenotypes (dbGaP)12 (accession number phs001739.v1.p1).
Additional associated metadata were requested and obtained from the RAMS Registry (https://ramsregistry.vcu.edu) (PRJNA430482) or from the senior author (Projects PRJEB11895, PRJEB12577, PRJEB21325, and PRJEB30642), or downloaded from the publication by the Kindinger et al.(ref) (PRJEB11895 and PRJEB12577).
Data for accession number phs001739.v1.p1 are exclusively available via dbGap after following the application procedures there.
The aggregated vaginal microbiome dataset can be downloaded from the March of Dimes Prematurity Research Database excluding the datasets that we are unable to share due to legal restrictions (SDY2187).

https://pretermbirthdb.org

https://immport.org

https://dbgap.ncbi.nlm.nih.gov/aa/wga.cgi

https://www.ncbi.nlm.nih.gov/sra

https://www.ebi.ac.uk/ena/browser

## Acknowledgements

We thank members of the Sirota Lab, University of California, San Francisco, for useful discussion. This study was supported by the March of Dimes (ALP, TTO, AR, AST, CWYH, RJW, GA, IK, SN, YSLL, PRB, DAM, SVL, DKS, NA, JLG, MS); Spanish Ministry of Science, Innovation and Universities through FPU program FPU18/0177; EST22/00170 (ALP), Instituto de Salud Carlos III (Spanish Ministry of Science and Innovation) through Miguel Servet program CP20/00118 and co-funded by European Union (PGD), NIH grants R35GM138353 (NA), 1R01HL139844 (NA), 3P30AG066515 (NA), 1R61NS114926 (NA), 1R01AG058417 (NA), R01HD105256 (NA, MS), and P01HD106414 (NA); the Burroughs Welcome Fund (NA); the Alfred E. Mann Foundation (NA); and the Robertson foundation (NA).

## Author Contributions

JLG and MS conceived the study. CWYH, RJW, and ALT generated and shared data for the validation dataset. JLG, TTO, AST, AR, and SSM aggregated the training datasets. JG and AR normalized the training and validation datasets. ALP and JG developed the application. TTO, JLG, and MS were major contributors in writing the manuscript. All authors read and approved the final manuscript.

## Ethics Statement

This work was approved by the National Heart, Lung, and Blood Institute (NHLBI) Clinical Data Science Institutional Review Board (CDS-IRB) in study number 2021-040, and reliance was granted to the NHLBI CDS-IRB by the University of California, San Francisco Institutional Review Board in study number 21-35274.

## References

1. Hyman, R. W. et al. Diversity of the Vaginal Microbiome Correlates With Preterm Birth. Reprod. Sci. 21, 32–40 (2014).

2. DiGiulio, D. B. et al. Temporal and spatial variation of the human microbiota during pregnancy. Proc. Natl. Acad. Sci. U. S. A. 112, 11060–11065 (2015).

3. Callahan, B. J. et al. Replication and refinement of a vaginal microbial signature of preterm birth in two racially distinct cohorts of US women. Proc. Natl. Acad. Sci. U. S. A. 114, 9966–9971 (2017).

4. Kindinger, L. M. et al. The interaction between vaginal microbiota, cervical length, and vaginal progesterone treatment for preterm birth risk. Microbiome 5, 6 (2017).

5. Kindinger, L. M. et al. Relationship between vaginal microbial dysbiosis, inflammation, and pregnancy outcomes in cervical cerclage. Sci. Transl. Med. 8, 350ra102 (2016).

6. Brown, R. G. et al. Vaginal dysbiosis increases risk of preterm fetal membrane rupture, neonatal sepsis and is exacerbated by erythromycin. BMC Med. 16, 9 (2018).

7. Brown, R. G. et al. Establishment of vaginal microbiota composition in early pregnancy and its association with subsequent preterm prelabor rupture of the fetal membranes. Transl. Res. J. Lab. Clin. Med. 207, 30–43 (2019).

8. Caporaso, J. G. et al. Global patterns of 16S rRNA diversity at a depth of millions of sequences per sample. Proc. Natl. Acad. Sci. 108, 4516–4522 (2011).

9. Schmidt, T. S. B., Matias Rodrigues, J. F. & von Mering, C. Limits to robustness and reproducibility in the demarcation of operational taxonomic units. Environ. Microbiol. 17, 1689–1706 (2015).

10. Sirota, M. et al. Enabling precision medicine in neonatology, an integrated repository for preterm birth research. Sci. Data 5, 180219 (2018).

11. Bhattacharya, S. et al. ImmPort: disseminating data to the public for the future of immunology. Immunol. Res. 58, 234–239 (2014).

12. Mailman, M. D. et al. The NCBI dbGaP database of genotypes and phenotypes. Nat. Genet. 39, 1181–1186 (2007).

13. Leinonen, R., Sugawara, H., Shumway, M., & International Nucleotide Sequence Database Collaboration. The sequence read archive. Nucleic Acids Res. 39, D19–21 (2011).

14. Leinonen, R. et al. The European Nucleotide Archive. Nucleic Acids Res. 39, D28–D31 (2011).

15. Golob, J. L. et al. Microbiome Preterm Birth DREAM Challenge: Crowdsourcing Machine Learning Approaches to Advance Preterm Birth Research. 2023.03.07.23286920 Preprint at https://doi.org/10.1101/2023.03.07.23286920 (2023).

16. Minot, S. S. et al. Robust Harmonization of Microbiome Studies by Phylogenetic Scaffolding with MaLiAmPi. 2022.07.26.501561 Preprint at https://doi.org/10.1101/2022.07.26.501561 (2022).

17. France, M. T. et al. VALENCIA: a nearest centroid classification method for vaginal microbial communities based on composition. Microbiome 8, 166 (2020).

